# Early Experience with Ivabradine for Focal Atrial Tachycardia in Pediatric Patients with Congenital Heart Disease

**DOI:** 10.1101/2023.06.07.23291097

**Authors:** Drishti Tolani, Nawin L. Ramdat Misier, Manal Alqahtani, Kaitlin Tindel, William A. Scott, Hoang H. Nguyen

## Abstract

**Background:** Ivabradine is a promising anti-arrhythmic therapy for automatic arrhythmias such as inappropriate sinus tachycardia, junctional ectopic tachycardia, and focal atrial tachycardia (AT). However, experience with ivabradine in pediatric patients, especially those with congenital heart disease (CHD) and focal AT, remains limited. We report our findings using ivabradine for focal AT in infants and children with CHD to assess its efficacy and safety.

**Method:** A retrospective analysis was conducted on all pediatric patients (<21 years) diagnosed with CHD at Children’s Health of Dallas, who were treated with ivabradine for focal AT. Patient demographics, arrhythmia diagnosis, anti-arrhythmic therapies, and adverse effects were evaluated. A positive response was defined as complete rhythm control within 24 hours of initiation of ivabradine.

**Results:** Fifteen patients (median age 7 [1-8] months; 9 males (60%)) were included in this study, including 12 (80%) complex CHD. Fourteen patients (93%) had unifocal AT and one (7%) had multifocal AT. The AT occurred in the early post-operative period in six patients (40%). In two patients (13%) ivabradine was used as monotherapy. Positive response to ivabradine was observed in 12 patients (80%). Adverse events occurred in 7 patients (47%) consisting of bradycardic, which was transient, or resolved upon reducing the ivabradine dosage.

**Conclusion:** In infants and children with CHD, ivabradine was efficacious for the treatment of focal AT without major complications. Bradycardia is a frequent adverse event. Therefore, close monitoring may be required during initiation of therapy.

## Introduction

Ivabradine is an anti-arrhythmic agent that reduces the heart rate by inhibiting the pacemaker current, which is responsible for spontaneous depolarization of the sinoatrial node.(1) Currently, ivabradine is primarily used for treatment of inappropriate sinus tachycardia and sinus tachycardia related to heart failure, in adults.(2, 3) More recently, there has been increasing experience with ivabradine for junctional ectopic tachycardia (JET) in pediatric patients.(4) As abnormal spontaneous diastolic depolarization is the pathophysiologic mechanism for focal atrial tachycardia (AT), i ivabradine may be an effective therapy.(5)

Experience with ivabradine in pediatric patients with focal AT is limited to small case series, often lacking patients with congenital heart disease (CHD).(6-11) In young patients with CHD, AT is a significant source of morbidity and mortality.(12, 13) Multiple anti-arrhythmia agents are moderately effective in treating AT; however, all have potentially significant side effects, which could outweigh their benefits.(12-14) An important advantage of ivabradine is its high selectivity. It does not have negative inotropic effects or prolong repolarization.(1, 4, 15)

We, therefore, report in this single-center study our early experience using ivabradine for focal AT in infants and children with CHD to assess its efficacy and safety.

## Methods

A retrospective review was conducted on all pediatric patients at Children’s Health of Dallas who were treated with ivabradine for focal AT. The inclusion criteria for the study were pediatric patients <18 years with CHD and focal AT, who were treated with ivabradine as inpatients.

Our center has been using ivabradine for AT since August 2020, and all retrospective data available from August 2020 until May 2023 were collected. Local institutional review board approval was obtained, and informed consent was waived.

### Ivabradine protocol

Data were retrieved from medical records and included demographic characteristics, type of CHD, date and types of cardiac procedures, comorbidities, pharmacological therapy before ivabradine treatment, electrocardiographic recordings, and echocardiographic recordings.

Analysis of inpatient telemetry, standard 12 lead electrocardiogram and atrial wire studies were utilized for diagnosis. Focal AT was confirmed by a pediatric electrophysiologist. Incessant focal AT was defined as a continuous tachycardia without spontaneous termination or continuous paroxysms of tachycardia separated by ≤2 sinus beats and lasting >24 hours.

Pediatric patients were treated with ivabradine when able to receive PO medication. Decision to initiate ivabradine was left to the treating electrophysiologist in consultation with the primary care team. In general, first-line treatment with ivabradine was considered when the AT was not critical. Ivabradine was also used as second or third-line agent to supplement or replace medications that were not completely effective. Generally, ivabradine was administered at an initial dose of 0.05 mg/kg (a dosage of 2.5 mg for patients weighing⍰>⍰30 kg) every 12 hours under continuous telemetry of ECG and oxygen saturation. Blood pressure was monitored by indwelling line or frequent noninvasive measurements. Treatment was considered successful if rhythm control was achieved within 24 hours of ivabradine initiation. The dosage of ivabradine was increased to 0.1 mg/kg, when the first dose showed no effect on the heart rhythm or rate. If ivabradine was not successful within 24 hours, it was discontinued. In patients for whom first-line ivabradine was ineffective, other antiarrhythmic drugs were prescribed. In cases of successful ivabradine therapy, ongoing discussion with the primary care time determined the duration of therapy. Ivabradine dosage was lowered for clinically relevant bradycardia. Side effects, including acute hemodynamic changes, bradycardia and pro-arrhythmia, were specifically sought out.

### Statistical analysis

Continuous data were described as medians [ranges] and categorical variables were described as counts and percentages. Estimated statistics were performed for statistical comparison.(16)

## Results

### Study population

Fifteen patients (9 males (60%)) were included in this study. The median age at ivabradine initiation was 7 [1-8] months, with 6 patients (40%) being younger than 3 months old. Of the 12 the patients with complex CHD, 8 patients had single ventricle physiology and six patients had heterotaxy syndrome. Four patients (27%) had diminished systolic ventricular function. Baseline characteristics of the study are summarized in **Table 1**.

**Table 1.**

|  | Study population (n=15) |
| --- | --- |
| **Baseline characteristics** |  |
| Age (months) | 7 [1-18] |
| Weight (kg) | 6.7 [3.9-8.5] |
| Gender (kg) | 9/15 (60%) |
| Complex congenital heart disease | 12/15 (80%) |
| Diminished systolic ventricular function | 4/15 (27%) |
| **Medication characteristic** |  |
| Therapy indication: |  |
| First line | 4/15 (27%) |
| Second line | 6/15 (40%) |
| Third line | 1/15 (7%) |
| Breakthrough | 4/15 (27%) |
| Maximum ivabradine dose (mg/kg/dose) | 0.07 [0.05-0.10] |
| Monotherapy | 2/15 (13%) |
| Concurrent anti-arrhythmic medications: |  |
| Flecainide | 4/15 (27%) |
| Amiodarone | 4/15 (27%) |
| Beta-blocker | 4/15 (27%) |
| Digoxin | 3/15 (20%) |
| Dexmedetomidine | 2/15 (13%) |
| **Outcomes** |  |
| Acute success | 12/15 (80%) |
| Adverse events: | 7/15 (33%) |
| Sinus bradycardia | 5/15 (33%) |
| Functional bradycardia | 2/15 (13%) |

Fourteen patients (93%) had unifocal ectopy AT and one patient (7%) had multifocal AT. In total, six patients (40%) developed AT early after cardiac surgery or interventional catheterization. Six out of 15 ATs were incessant. None of the patients had prior atrial or ventricular arrhythmias.

### Ivabradine regimen

Ivabradine was used as first-line therapy in 4 patients (27%), second-line therapy in 6 (40%) and third line in 1 (7%) (**Table 1**). In 4 additional patients (27%), ivabradine was administrated to avoid breakthrough of ATs. In total, 13 patients (87%) used ivabradine in combination with other anti-arrhythmic medications, while only a minority (13%) used it as monotherapy. Ten patients (67%) were on a combination of ivabradine and one other anti-arrhythmic agent, including beta-blocker (N=2), flecainide (N=2), dexmedetomidine (N=2), amiodarone (N=2) and digoxin (N=2). Three patients (20%) were on a combination with ≥2 anti-arrhythmic drugs: ivabradine with flecainide and amiodarone; ivabradine with a beta-blocker and amiodarone; or ivabradine with a beta-blocker, flecainide, and digoxin.

The initial ivabradine dosage was 0.05 [0.05-0.05; min 0.02 – max 0.1) mg/kg. Two patients received adult dosing at 2.5 mg (34 kg) and 5 mg (70 kg), resulting in 0.07 mg/kg in both respectively. The maximum dosage during treatment was 0.07 [0.05-0.1; min 0.038-max 0.157) mg/kg, which was higher than the initial dose in 7 patients (all <1 year of age).

### Efficacy outcomes

Positive response to ivabradine was observed in 12 patients (80%). During their in-hospital stay, they were treated with ivabradine for a median duration of 6 [5-26] days, and none had recurrence while on ivabradine therapy. In 3 patients (20%), all with post-operative AT, ivabradine was unsuccessful in providing adequate rhythm control. The first patient had incessant AT which did not respond to ivabradine despite optimization of the dose. After discontinuation of ivabradine, rhythm control was achieved with after initiation of multiple other anti-arrhythmic agents. The second patient continued to have multifocal AT on ivabradine, although a more organized rhythm was observed. The third patient remained to have breakthrough focal AT on ivabradine. After addition of flecainide, the patient was free of AT.

After discharge, 3 of the 12 responders (25%) no longer required treatment. Two patients (13%) had mono-drug therapy with ivabradine after discharge. The first patient was initially weaned off ivabradine after discharge, but had recurrent AT 1 year after discharge, which responded to ivabradine. The second patient received maintenance therapy with ivabradine, without recurrence during follow-up. The 7 other patients (58%) either continued using one or multiple other antiarrhythmic drugs, without ivabradine.

### Safety outcomes

In total, 7 patients experienced adverse events (47%), all consisting of bradycardia alone. Bradycardia occurred at a median of 3 [IQR 1-3] days after initiation of ivabradine, with a maximum of 7 days. Five patients had sinus bradycardia, and 2 had functional bradycardia due to blocked premature atrial contractions (PACs). All patients with bradycardia were receiving concomitant anti-arrhythmic agents. Using estimated statistics, the unpaired median differences in age between patients with and without bradycardia was -3.5 [95.0%CI -85.0, 7.5] months (two-sided permutation t-test P value = 0.492).

In 4 patients, either a single dose was skipped or the chronic dose was halved, which successfully restored sinus rhythm within 12 hours. In 1 patient, ivabradine was discontinued due to bradycardia after conversion to sinus rhythm, after which patient had recurrence of AT. Flecainide and propranolol successfully terminated the AT, but the AT recurred once these medications were discontinued. Subsequently, ivabradine was initiated again and successfully converted the AT to sinus rhythm without AT or bradycardia recurrence. In another patient ivabradine was replaced by flecainide which resolved the functional bradycardia. In the remaining 2 patients ivabradine regimen was not changed during well tolerated bradycardia that spontaneously resolved later the same day.

## Discussion

### Key findings

In this retrospective single center study, we assessed for the first time the efficacy and safety of ivabradine for focal AT in infants and children with CHD. The 80% response rate was high, although often in combination with other anti-arrhythmic agents. Adverse events were limited to bradycardia without significant clinical consequences.

### Clinical experience with ivabradine

In recent years, additional indications for ivabradine have been explored, particularly in pediatric patients, due to the favorable side effect profile.(4, 6, 15) In these young patients, Ivabradine has recently been shown to be a promising pharmacological agent in the treatment of heart failure and JET.(15) Similar to the adult population, there has also been increasing interest in its use for automatic AT. However, the experience was limited to several case-reports and small series in pediatrics, often with normal cardiac anatomy.(6-10) Recently, Xu et all reported its single center experience with ivabradine monotherapy in 12 pediatric patients with focal AT and no CHD, who were resistant to conventional anti-arrhythmic agents.(6) Ivabradine was effective in 50% of the patients, and well tolerated without any events of bradycardia. However, except for a case-report, outcomes of ivabradine in pediatric with congenital heart disease were unknown.(9)

Focal AT are not uncommon in pediatric patients with congenital heart disease, especially in the early post-operative period.(14) In these patients, AT is often associated with hemodynamic comprise and a three times higher mortality rate.(13) Not only are the current anti-arrhythmogenic agents characterized by suboptimal efficacy, they also have negative inotropic effects, potential for toxicity, and carry increased risks for other arrhythmias.(12-14) In this vulnerable population, catheter ablation is often unattractive due to technical complexity, moderate success rates and risk for complications.(12, 14) We now demonstrate in a sizable population, that ivabradine is effective for treatment of focal AT in pediatric patients with CHD, often when other anti-arrhythmic agents failed. Moreover, its efficacy may be higher compared to pediatric patients (6/12, 50%) and adult patients with normal anatomy (18/34, 64%).(5, 6)

### Alternative mechanisms underlying focal AT

Although focal AT are from a mechanistic point of view due enhanced automaticity, it is not restricted to this mechanism.(17) Micro-reentry and triggered activity may also underly focal AT, which could explain the non-response in selected patients with focal AT. Various reports show that these alternative mechanisms are frequent in pediatric and adult patients with and without congenital heart disease.(17-19) In addition, increased automaticity – and thus pacemaker activity – are not solely dependent on HCN channels, which are the electrical target for ivabradine treatment. Enhanced function of other ionic channels and their currents may also be responsible for spontaneous diastolic depolarizations in selected patients with automatic FAT, rendering them insensitive to ivabradine.(1)

### Bradycardia and side effects

The minimal side-effect profile of ivabradine has been a major reason for the increased interest in ivabradine as an alternative anti-arrhythmic agent for focal AT.(1, 4, 6, 15) Unlike traditional anti-arrhythmic agents, ivabradine has minimal effects on blood pressure and inotropy. However, an important adverse effect is bradycardia.(15, 20) While there were no documented episodes of bradycardia in the case-series from Xu et al., we observed bradycardia in 47% of patients.(6) Interestingly, bradycardia was not just due to depressed sinoatrial node pacemaker function, 2 out of 7 patients had functional bradycardia due to blocked PACs. Ivabradine can also inhibit the funny current in the atrioventricular node, resulting in functional bradycardia in presence of PACs.(21)

In pediatric patients with complex CHD, bradycardic events may be more significant as this population often has diminished ventricular function. In addition, pediatric patients < 1 year of age are especially dependent on chronotropic competence. Importantly, however, bradycardic events in the current study were transient or responded to reducing the ivabradine dosage. A recent report showed that in the case of ivabradine overdoses (0.5 mg/kg due to dosing error), bradycardia did not result in hemodynamic instability in a child with AT. In addition, within 28 hours of discontinuation of ivabradine sinus rhythm was returned.(11) Also, pediatric patients with early post-operative AT often have temporary atrial or ventricular pacing wires which could ensure sufficient ventricular rate in case of persisting bradycardia.

### Limitations

Although this is the first large retrospective single center report on the use of ivabradine in pediatric with CHD, there are several limitations inherent to its design. Due to its retrospective nature, there was no standardized protocol in initiation of ivabradine, resulting in first, second and third-line therapy regimens in combination with various other anti-arrhythmic agents. Future studies need to assess the efficacy and safety of monotherapy separately from interaction with specific other anti-arrhythmic agents. Due to the relatively small sample size, the study was unable to identify predictors for response, adverse events and determine the optimal dose for treatment.

## Conclusion

In infants and children with CHD, ivabradine showed a high efficacy for the treatment of focal AT without any major complications. Bradycardia is a frequent adverse event; although transient and short-lived, these events may require close monitoring especially in the post-operative course.

## Data Availability

All data produced in the present study are available upon reasonable request to the authors.

## Abbreviations

AT: atrial tachycardia
CHD: congenital heart disease
JET: junctional ectopic tachycardia

